# Neurophysiological and brain structural markers of cognitive frailty differs from Alzheimer’s disease

**DOI:** 10.1101/2021.01.06.21249338

**Authors:** Ece Kocagoncu, David Nesbitt, Tina Emery, Laura Hughes, Richard N. Henson, Cam-CAN, James B. Rowe

## Abstract

With increasing life span, there is growing importance of understanding the mechanisms of successful cognitive ageing. In contrast, cognitive frailty has been proposed to be a precursor to Alzheimer’s disease. Here we test the hypothesis that cognitively frail adults represent a branch of healthy ageing, distinct from latent dementia. We used electro-magnetoencephalography and magnetic resonance imaging to investigate the structural and neurophysiological features of cognitive frailty in relation to healthy aging, and clinical presentations of mild cognitive impairment and Alzheimer’s disease. Cognitive performance of the cognitively frail group was similar to those with mild cognitive impairment. We used a novel cross-modal oddball task to induce mismatch responses to unexpected stimuli. Both controls and cognitively frail showed stronger mismatch responses and larger temporal grey matter volume, compared to people with mild cognitive impairment and Alzheimer’s disease. Our results suggest that cognitively frail represents a spectrum of normal ageing rather than incipient or undiagnosed Alzheimer’s disease. Lower cognitive reserve, hearing impairment and medical comorbidity might contribute to the aetiology of cognitive impairment.

## 1. Introduction

With longer life span and an older population, there is a pressing need to understand the mechanisms that determine cognitive ageing, and its relationship to dementias. There is increased interest in a group of adults referred to here as “cognitively frail”, defined by reduced cognitive function in the absence of subjective memory complaints, or a clinical diagnosis diagnosis of dementia or precursor dementia state, or other pre-existing neurological explanation (Kelaiditi et al., 2013). Cognitive frailty has been linked to a higher risk of dementia, and is often seen as a precursor to Alzheimer’s disease (Buchman et al., 2007; Kojima et al., 2016; Panza et al., 2006; Shimada et al., 2018; Wang et al., 2017). In addition, cognitive frailty, irrespective of comorbid physical frailty, is associated with longitudinal decline in functional abilities, activities of daily living (Avila-Funes et al., 2009; Shimada et al., 2016), increased hospitalisation and all-cause mortality rate (Cano et al., 2012; Feng et al., 2017; Solfrizzi et al., 2012). Poor baseline cognitive performance is a predictor of future cognitive decline and all-cause mortality in long-term follow-up (Avila-Funes et al., 2012; Solfrizzi et al., 2017b).

However, there is an alternative hypothesis of cognitive frailty: poor cognitive performance may reflect accelerated or adverse aspects of normal ageing process, without incipient Alzheimer’s disease or other dementia. Psychosocial, behavioural factors and medical comorbidities may contribute to cognitive frailty in the absence of latent degenerative or vascular dementia pathologies. For example cognitively frail individuals are four times more likely to come from disadvantaged socioeconomic backgrounds, and twice as likely to have lower educational qualifications (Rogers et al., 2017). They are more likely to suffer from poorer nutrition (Chye et al., 2018; Mulero et al., 2011; Talegawkar et al., 2012), have lower levels of physical exercise (Landi et al., 2010; Rogers et al., 2017), and have more medical comorbidities such as cardiovascular disease (Fuhrmann et al., 2019; Langlois et al., 2012; Patrick et al., 2002), chronic inflammation (Cappola et al., 2003; Walston et al., 2002; Weaver et al., 2002) and hearing impairment (Panza et al., 2015a; Valentijn et al., 2005).

This study aimed to determine whether cognitive frailty had brain structural and physiological characteristics of normal ageing, or early Alzheimer’s disease. It is set in the context of the population-based cohort in the Cambridge Centre for Ageing and Neuroscience (Cam-CAN). We focus on the cognitive component of frailty, and use magnetic resonance imaging (MRI) and electro-magnetoencephalography (E/MEG) to investigate its structural and neurophysiological features. We compare people with cognitive frailty to those with a clinical diagnosis of mild cognitive impairment or Alzheimer’s disease. In Alzheimer’s disease amyloid beta plaques and neurofibrillary tau tangles form early in the entorhinal cortex and hippocampi, and later throughout the medial temporal lobe and connected structures (Braak et al., 2006; Hardy and Selkoe, 2002; Schöll et al., 2016), causing disruptions in synaptic and neural function (Hsieh et al., 2006; LaFerla and Oddo, 2005; Li et al., 2009; West and Bhugra, 2015), as clinical symptoms emerge. As a corollary, if cognitively frail adults are in the prodromal stage of Alzheimer’s disease, then one would expect similar structural changes in the hippocampus and temporal cortex, and similar physiological change.

We use E/MEG to capture synaptic dysfunction and disruptions in neural signalling. E/MEG has been shown to capture variations in cognitive function in healthy successful aging (Coquelet et al., 2017; Price et al., 2017; Tsvetanov et al., 2015; Vlahou et al., 2014), and early signatures of mild cognitive impairment and Alzheimer’s disease (de Haan et al., 2012; Hughes et al., 2019; Kocagoncu et al., 2020; Maestú et al., 2015; Osipova et al., 2005; Stam et al., 2006) (Koffie et al., 2011; LaFerla and Oddo, 2005).

To assess hippocampal-dependent functions, we designed a novel task called the *cross-modal oddball task*. This is based on into the critical role of the hippocampus in associative memory (Chua et al., 2007; Giovanello et al., 2004; Konkel et al., 2008; Köhler et al., 2005). The task consists of a rapid series of trials, comprised of an image paired with a sound. The mismatch responses arise from pairs that include either a novel sound (i.e. *novelty deviant*), or a sound that is not novel, but was previously associated with a different image (i.e. *associative deviant*). Hippocampal dysfunction affects the ability to establish cross-modal associations and therefore attenuates the associative deviant response. The novelty deviant is akin to the classic mismatch negativity response, an index of auditory predictive coding. The classic mismatch negativity is not hippocampal dependent, but is abnormal Alzheimer’s disease (Jiang et al., 2017; Laptinskaya et al., 2018; Pekkonen et al., 2001; Ruzzoli et al., 2016). Although neurophysiological responses in the hippocampus are difficult to detect with E/MEG, owing to its depth and orientation, a strong mismatch response can be recorded from auditory cortex where sensory predictions are established from cross-model associative learning. We therefore focus on the mismatch response in the auditory and frontal cortices, which are regions activated in conventional auditory oddball paradigms (Garrido et al., 2009; Hughes et al., 2018; Hughes and Rowe, 2013; Pekkonen, 2000; Phillips et al., 2016).

We compared cognitively frail adults with cognitively normal healthy controls and two groups of patients from regional memory clinics, with Alzheimer’s disease and mild cognitive impairment. We proposed that if cognitive frailty represented a spectrum of normal ageing, rather than latent disease, then the neurophysiological responses and structural features of cognitively frail adults would resemble the cognitively healthy adults rather than the patients with mild cognitive impairment or Alzheimer’s disease.

## 2. Materials and Methods

### 2.1. Cam-CAN

The Cam-CAN Frail Project is an extension of the large-scale cross-sectional population-based Cam-CAN study, examining the sub-population of cognitively frail adults identified from home screening visits (Shafto et al., 2014). The Cam-CAN Frail protocol comprised three sessions. First, a visit to the participant’s home to assess lifestyle, health and cognitive performance on an extensive neuropsychological test battery. The battery included the revised Addenbrooke’s Cognitive Examination (ACER), Mini Mental State Examinations (MMSE), Wechsler Adult Intelligence Scale logical memory test, Spot the Word test, simple choice reaction time, famous faces test, four-mountains task, virtual object location and orientation, Rey figure recall, and the trail making test. In the second session, participants underwent E/MEG scanning and completed the Cattell and digit symbol tests. During the E/MEG recording, participants completed the cross-modal oddball task. In the final session, participants had a functional and structural magnetic resonance imaging and completed the Hotel task. The study was approved by the East of England – Cambridge Central Research Ethics Committee (10/H0308/50).

### 2.2. Participants

Participants consisted of community-dwelling older healthy controls, cognitively frail, and patients diagnosed with either mild cognitive impairment or Alzheimer’s disease (**Table 1**). Participants were older than 50 years and were fluent speakers in English, with mental capacity to consent. Participants did not have any established neurological condition (other than mild cognitive impairment or Alzheimer’s disease in the patient groups), nor significant psychiatric illness. Healthy controls had not taken part in the “Cam-CAN 700” or “Cam-CAN 280” stages, but had scored >25/30 on MMSE or >88/100 on the ACER during the home interview. The Cam-CAN home visits acquired lifestyle and cardiovascular risk characteristics (alcohol and smoking, hypertension, history of stroke and heart attack). The cognitively frail adults scored below 25/30 on the MMSE and/or below 88/100 on the ACER. In addition, patients were recruited from local specialist memory clinics and had mild cognitive impairment or probable Alzheimer’s disease diagnosed as per the Petersen and McKhann criteria respectively (McKhann et al., 2011; Petersen et al., 2014). Eight of the MCI patients and two of the AD patients had positive cerebrospinal fluid biomarker status for Alzheimer’s disease. Six of the MCI patients had negative biomarker status. The biomarker status of the remaining MCI and Alzheimer’s disease patients was unknown.

**Table 1.** Sample characteristics

|  | Controls | Frail | MCI | Alzheimer |
| --- | --- | --- | --- | --- |
| Sample size | 38 (17 F) | 26 (14 F) | 15 (4 F) | 11 (6 F) |
| Age | 72.19 ± 8.88 | 79.98 ± 9.50 | 75.54 ± 7.60 | 74.53 ± 11.17 |
| Education (years) | 14.97 ± 3.86 | 11.07 ± 2.88 | 16.68 ± 4.99 | 11.55 ± 3.54 |
| Hearing left (dB) | 51.92 ± 13.32 | 44.15 ± 16.06 | 52.88 ± 14.77 | 54.80 ± 10.01 |
| Hearing right (dB) | 53.55 ± 12.44 | 43.38 ± 16.35 | 57.63 ± 9.40 | 50.00 ± 12.01 |
| MMSE (/30) | 28.34 ± 1.47 | 26.07 ± 2.28 | 26.37 ± 2.73 | 23.20 ± 2.78 |
| ACE-R (/100) | 93.71 ± 3.04 | 80.92 ± 6.01 | 83.68 ± 8.41 | 68.6 ± 8.11 |
| ACE-R memory (/26) | 23.63 ± 1.94 | 18.38 ± 3.69 | 16.81 ± 6.09 | 10.7 ± 3.43 |
| Training test (/100) | 65.78 ± 23.55 | 50.96 ± 20.59 | 44.16 ± 24.02 | 37.50 ± 27.95 |

### 2.3. E/MEG and MRI acquisition

E/MEG data were acquired using the Elekta Vector View system with 204 planar gradiometers and 102 magnetometers. Simultaneous EEG data were acquired using a 70-channel Easycap. Participants’ horizontal and vertical eye movements, and the cardiac activity were recorded using bipolar electro-oculogram and electro-cardiogram electrodes. Five head position indicator coils were placed on the EEG cap, to track the head position every 200 ms. For coregistration of the participant’s T1-weighted MRI scan to the MEG sensors, three fiducial points (nasion, left and right pre-auricular) and a minimum of 100 head shape points were digitized using Polhemus digitization.

Participants were seated in a magnetically shielded room (IMEDCO) and positioned under the MEG scanner. Auditory stimuli were delivered binaurally through MEG-compatible ER3A insert earphones (Etymotic Research). The delay in sound delivery due to the length of earphone tubes and sound card was 26 ± 2 ms on average. Visual stimuli were presented on the screen positioned 1.22 m in front of the participant’s visual field. Simultaneous E/MEG was recorded continuously at 1000 Hz with a high-pass filter of 0.03 Hz. Before the E/MEG recording, participants performed an automated hearing test in the MEG scanner, to make sure that the earphones were working properly. They were presented pure tones at the frequency of 1000 Hz to either ear with varying loudness. Participants were instructed to press the button when they heard the tone. The mean hearing levels of each group is given in **Table 1**, where the normal range is expected to fall within 45-75 dB.

T1-weighted structural images were acquired on a Siemens 3T Magnetom Prisma MRI Scanner using a MPRAGE sequence (repetition time = 2250ms, echo time = 2.99ms; inversion time = 900ms; flip angle= 9 degrees; field of view = 256mm x 240mm x 192mm; voxel size = 1mm isotropic; GRAPPA acceleration factor = 2; acquisition time = 4 minutes 32 seconds). Four participants did not tolerate MRI due to claustrophobia.

### 2.4. Stimuli

The stimuli consisted of abstract images and pure tones. There were four images with distinct patterns. The tones had the following frequencies: 503 Hz, 719 Hz, 1021 Hz and 1451 Hz. Harmonic tones were avoided by choosing frequencies of prime numbers and varying them by at least 3 semi-tones. There were four types of trials. 1) *Standard (STD) trials* were image-tone pairs that participants trained on prior to the task. Standard pairs were the trials presented most frequently. 2) *Associative deviant (DA) trials* presented the same images of the standard pairs but by shuffling the sounds. The DA trials were expected to capture the binding effect arising from a mismatch in association. 3) *Novelty deviant (DN) trials* presented the standard images with rare deviant tones. The frequencies used for the novel deviants were 599 Hz, 857 Hz, 1017 Hz, 1733 Hz. The DN trials were expected to capture the novelty effect, and were essentially the deviants used in conventional mismatch paradigms. The deviant trials were expected to induce a mismatch response with respect to the response to standard trials. 4) *Target trials*: The standard pairs, where the image was bound by a red circle. Target trials were included to make sure participants were attending to the stimuli. There were in total of 1000 standard trials and associative deviant, novel deviant and target trials were presented 48 times each. Therefore, the associative deviant, novel deviant and the targets were each encountered 4% of the time each, whereas standards 88% of the time.

### 2.5. Paradigm

The cross-modal oddball paradigm depends on both change detection and associative binding. This has two advantages. First, as MEG recording has lower signal to noise ratio in the subcortical areas and deeper sources compared to signal coming from superficial cortices (Goldenholz et al., 2009), the task was specifically designed to capture the indirect response in the superior temporal gyri and inferior frontal gyri, that are dependent on hippocampal associative learning. Secondly, the integrity of the auditory and frontal cortex is preserved until late stages of Alzheimer’s disease, allowing us to control for atrophy of the cortical generators of the mismatch response. The task was easy to perform both by all participant groups, require minimal training (reducing potential confounds such as education and cognitive strategies on performance).

Images were presented centrally on a grey screen bounded by a black circle for 800 ms. Then, 300 ms after image onset, the tone was played for 500 ms (**Figure 1A**). The lag was introduced to allow participants to form predictions about the upcoming auditory stimuli. In between trials, a black fixation square was presented for a jittered period of 300-500 ms, resulting in a stimulus onset asynchrony between 1000-1200 ms. E-Prime 2 (Psychology Software Tools) was used to present the stimuli and send triggers to the scanner.

**Figure 1.**
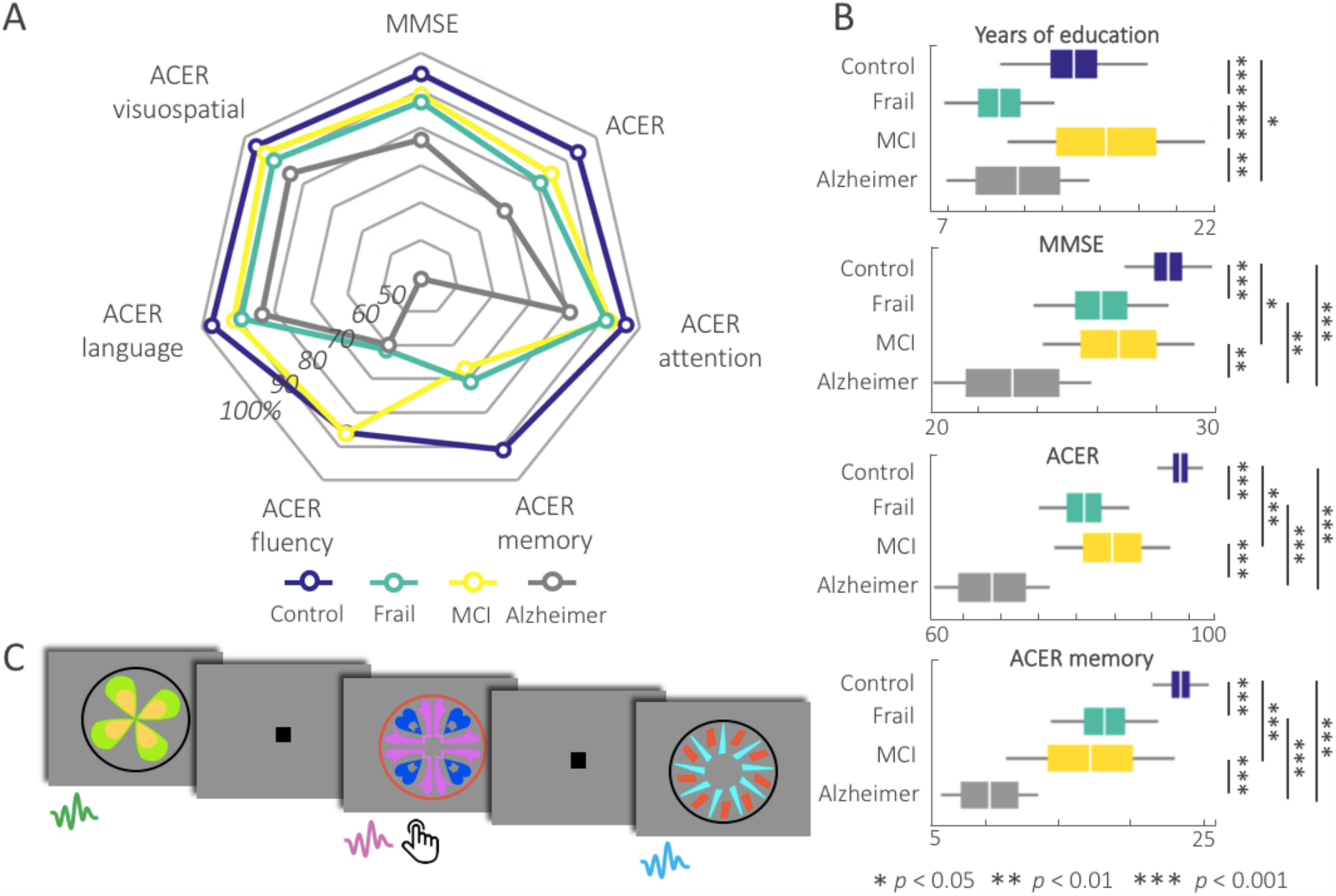
**A.** The radar chart displays the group means for the neuropsychological tests converted to percentages against the maximum score in each test for ease of comparison across groups. Note that the performance of the cognitively frail overlaps with the MCI across all tests except for ACER fluency. **B**. Group differences in education levels and neuropsychological tests. The cognitively frail had lower education levels than the controls. On the neuropsychological tests, the cognitively frail performed similar to the MCI group. **C**. Example stimuli from the cross-modal oddball task. The images were presented together with paired sounds after the 300 ms lag. Participants were asked to press the button whenever they saw a red circle around the image.

In the training phase, participants were presented in total four images and four tones (i.e. standard pairs), 25 times each, and were instructed to try to remember the pairings between the images and the tones. After the training, participants performed a short test where they listened to the four tones twice in a randomised order. After each tone, they were shown four images (i.e. chance level of %25) on the screen and were asked to select the image that was paired with that tone. Irrespective of the participant’s performance, training was repeated only once. Following the training participants moved on to the main task. Trials were presented in a different randomised order for each participant across four 5-minute long blocks. Participants were instructed to pay attention to the images and press the button with their right index finger when the image was bound by a red circle.

### 2.6. E/MEG pre-processing and source localization

The raw E/MEG data were pre-processed using MaxFilter 2.2.12 (Elekta Oy). Maxfiltering included detection and interpolation of bad sensors, signal space separation to remove external noise from the data and head movement correction. Cardiac and blink artefacts were removed using an independent component analysis with 800 maximum steps and 64 principal components via the EEGLAB toolbox (Delorme and Makeig, 2004). On average 2.38 blink components (*SD =* 0.58) and 1.30 cardiac components (*SD =* 0.49) were removed.

Data were further processed in SPM12 (www.fil.ion.ucl.ac.uk/spm). Data were bandpass filtered between 0-40 Hz using a fifth-order Butterworth filter. The continuous data were epoched between - 100 to 500 ms from the sound onset, and corrected for the auditory delay due to equipment. OSL’s artefact rejection algorithm (github.com/OHBA-analysis/osl-core) was used to remove any remaining artefacts (e.g. motor). Bad channels and trials marked by the algorithm were removed. On average 53.04 (*SD =* 35.88) trials and 10.76 channels (*SD =* 5.85) were removed per participant. Trials were averaged within condition, using robust averaging. Low-pass filter was re-applied to correct for the high-frequency noise introduced by robust averaging.

The E/MEG data were source localised using magnetometers, gradiometers and EEG (Henson et al., 2009). The source space was modelled with a medium sized cortical mesh consisting of 8196 vertices via inverse normalization of SPM’s canonical meshes. Sensor positions were coregistered to the native T1-weighted MPRAGE scans using the fiducial and head shape points after removing digitisation points around the nose. SPM’s canonical template brain was used for participants who did not tolerate the MRI scan. Single shell and Boundary Element models were used for forward modelling of MEG and EEG data respectively. Evoked signal was estimated over the trials using the COH solution in SPM which incorporates the minimum norm solution and a smooth source covariance matrix. All inversion accuracies were above 80% as measured by the R^2^ (*M =* 93.62; *SD =* 3.63).

The source localised data was extracted from 6 areas taken from the Automated Anatomical Labelling atlas: Heschl’s gyri, superior temporal gyri and inferior frontal gyri bilaterally. The ROI masks were resliced to 1mm isotropic thickness to allow maximum data extraction. For each participant and condition, the data were extracted from the peak within all the vertices that constitute each ROI. This is to maximise the signal to noise ratio in the data, and to account for individual variability in source activity. We had two contrasts of interest in the analyses. STD-DN contrast that captures the novelty mismatch effect, and the STD-DA contrast that captures the associative mismatch effect.

### 2.7. MRI pre-processing and grey matter analysis

The T1 image was rigid-body co-registered to a Montreal Neurological Institute (MNI) template and then corrected for image inhomogeneity and segmented into 6 tissue classes (grey matter, white matter, cerebrospinal fluid, bone, soft tissue, and residual noise) using SPM’s unified segmentation algorithm (Ashburner and Friston, 2005). The native space grey and white matter images for all participants were then submitted to diffeomorphic registration (DARTEL) (Ashburner, 2007) to create group template images. The group template was then normalised to the MNI template via an affine transformation and the combined normalisation parameters (native to group template and group template to MNI template) were applied to each individual participant’s grey matter image, including modulation in order to preserve local volume. Region of Interest (ROI) from the Harvard-Oxford atlas were then used to extract mean regional GMV from the bilateral hippocampal and entorhinal ROIs for each participant. The GMVs were compared across groups using ANCOVAs where age and total intracranial volume (TIV) were set as covariates.

To calculate local grey matter atrophy at the whole brain level we used voxel-based morphometry (VBM). Grey matter segments were thresholded with an absolute masking level of 0.1, and were smoothed with a Gaussian kernel at 8 mm full width half maximum. Grey matter volumes were compared across groups in pairwise t-contrasts in general linear models accounting for differences in age and total intracranial volume. The cluster level p-values were corrected for multiple comparisons using the family-wise error after a cluster defining threshold of *p* < 0.05.

### 2.8. RMS and statistical analyses

To investigate differences in time series, the root-mean-square of the time series at each ROI and trial were smoothed using a moving average at every 50 time points, to remove jumps. The RMS at each time point were then modelled using general linear models (GLM) accounting for differences in age and hearing levels, and tested for within group task effects by using t-contrasts. The contrasts compared the signal intensity between the DA-STD and DN-STD. The tests comparing the deviant effects were performed firstly within each participant group, to reveal task-specific effects. Secondly, these differences were tested across groups to test for interaction effects between conditions and groups. The observed cluster masses in the GLMs were corrected for multiple comparisons using permutation cluster statistics, by bootstrapping the design matrix using 1000 permutations at *p* = 0.05. The mean of the time series within each contrast were calculated for each participant within the 200-500 ms time window after removing outliers. This time window was selected because task effects were strongest after the N100. The linear relationship between these metrics and predictor variables were further tested through general linear models across the sample including age as a covariate, and after removing outliers. The predictors of interest were years of education, ACER total and memory subscale scores, and hippocampal and entorhinal grey matter volumes.

## 3. Results

### 3.1. Sample characteristics

Sample characteristics and scores on neuropsychological tests were compared across the groups using ANOVAs. Age (*F*(3,87) = 3.82; *p* = 0.012) and years of education (*F*(3,87) = 11.34; *p* < 0.001) were significantly different across groups. Tukey’s HSD tests showed that the cognitively frail group was older than the controls (*p* = 0.006). Controls’ education level was higher than the cognitively frail (*p <* 0.001), and the Alzheimer’s disease group (*p =* 0.032). Similarly, the MCI group had higher education level than the cognitively frail (*p <* 0.001) and the Alzheimer’s disease (*p =* 0.001) groups.

Hearing levels were tested for group differences both using ANOVAs and ANCOVAs to control for differences in age. There were no significant differences in hearing in the left ear. In the right ear there was a main group effect. The hearing levels (*F*(3,86) = 4.70; *p* = 0.004) of the cognitively frail were lower compared to both controls (*p* = 0.017) and the MCI (*p* = 0.005). When controlled for the differences in age (*F*(3,85) = 3.02; *p* = 0.034), the hearing levels on the right were still lower in the cognitively frail group compared to the MCI group (*p* = 0.019).

Chi-square tests compared the prevalence of lifestyle and cardiovascular risk factors between control and the cognitively frail groups (**Table S1**). The prevalence of daily drinking was significantly lower (*X*^*2*^(1) = 6.11; *p =* 0.006) in the cognitively frail group (19%) compared to controls (43%). The prevalence of hypertension was significantly higher (*X*^*2*^(1) = 2.74; *p =* 0.048) in the cognitively frail group (52%) compared to the controls (35%). There were no significant differences between the groups in the prevalence of smoking, history of stroke or heart attack.

### 3.1. Cognitive results

Cognitive scores were tested for group differences after controlling for differences in age (**Table 1**). The MMSE (*F*(3,86) = 17.64; *p* < 0.001), ACER total score (*F*(3,86) = 55.41; *p* < 0.001), ACER’s subscales in memory (*F*(3,86) = 37.35; *p* < 0.001), attention (*F*(3,86) = 9.05; *p* < 0.001), fluency (*F*(3,86) = 13.87; *p* < 0.001), language (*F*(3,86) = 7.90; *p* < 0.001) and visuospatial skills (*F*(3,86) = 11.15; *p* < 0.001) showed strong differences across the groups (**Figure 1A**). Results of the pairwise post-hoc comparisons are given in **Figure 1B** and **Table S2**. The cognitively frail performed similarly to the MCI group across all cognitive tests, except for the fluency subscale, where their scores were significantly lower than the MCI group (*p* < 0.001). All four groups performed above chance level on the training test. The scores were significantly different across groups (*F*(3,86) = 5.60; *p* = 0.001). Post-hoc comparisons showed that the controls performed significantly better than the MCI (*p =* 0.015) and Alzheimer’s disease (*p =* 0.006) groups. There were no significant differences between the training scores of the cognitively frail and the remaining groups.

### 3.2. Grey matter atrophy

Mean hippocampal GMV, entorhinal GMV, total GMV and total intracranial volume (TIV) were compared across the groups, corrected for age and TIV using ANCOVA. There were no significant differences between groups for TIV or total GMV. However, hippocampal (*F*(3,86) = 10.35; *p* < 0.001) and entorhinal (*F*(3,86) = 7.62; *p* < 0.001) GMVs showed a main group effect (**Figure 2A-B**). The hippocampal GMV in the control group was significantly larger compared to the MCI (*p <* 0.001) and Alzheimer’s disease groups (*p <* 0.001). Similarly, the entorhinal GMV of the control group was larger compared to the MCI (*p =* 0.001) and Alzheimer’s disease groups (*p =* 0.003). None of the comparisons showed significant differences between control and the cognitively frail groups.

**Figure 2.**
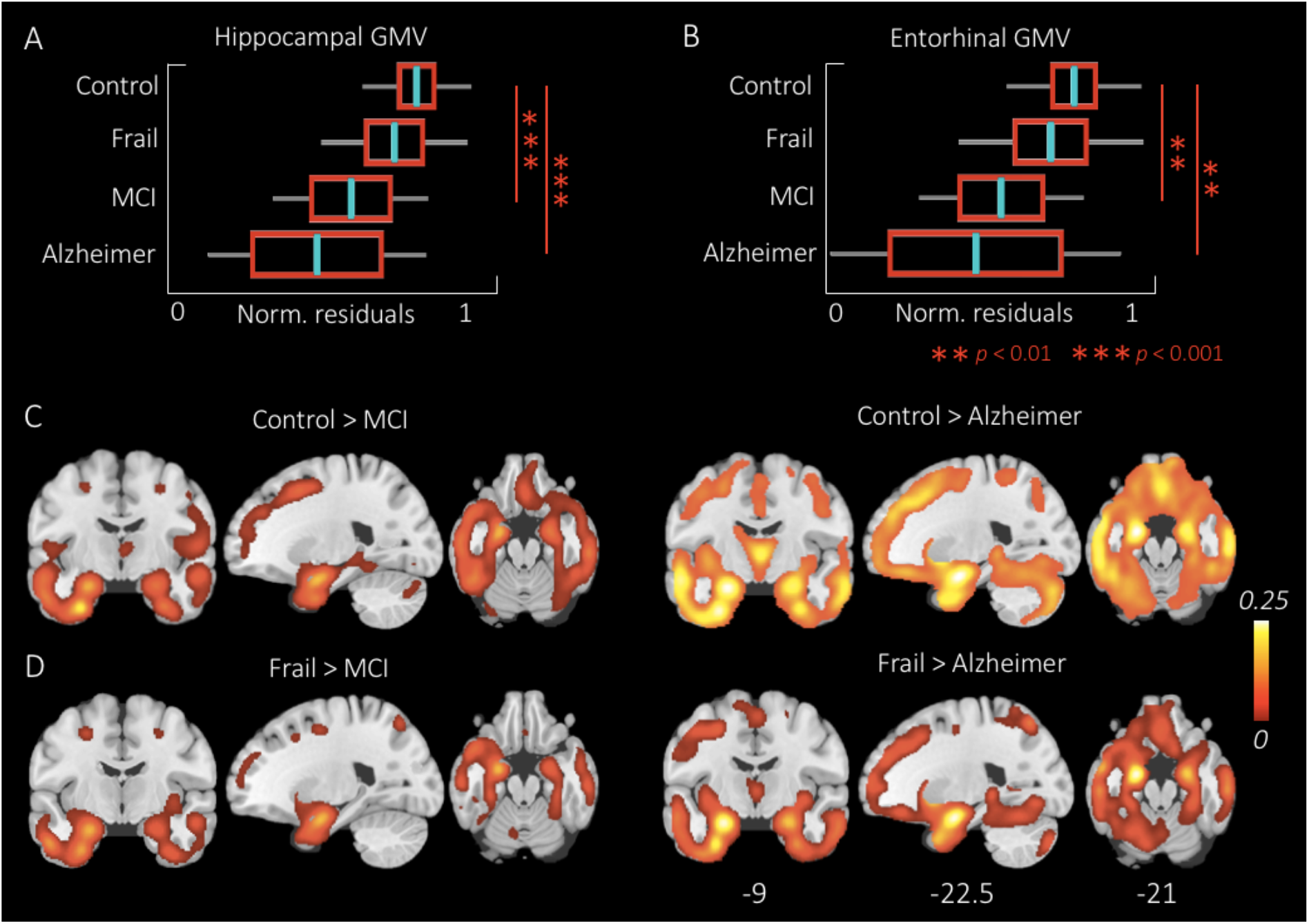
Grey matter analysis results. **A-B**. GMV differences across groups in the hippocampus and entorhinal cortex. The boxplots display the normalised residuals after correcting for differences in age and total intracranial volume (TIV). There were no significant differences in volume between the cognitively frail and the control group. **C-D**. The contrast images from the VBM analysis. Control and the cognitively frail show similar patterns of grey matter volume compared to the MCI and Alzheimer’s disease groups.

Atrophy was tested at the voxel level, using VBM (**Table 2**). There were no significant differences between the control and cognitively frail and between MCI and Alzheimer’s disease groups. As expected, the control group had significantly higher GMV in bilateral temporal cortices and hippocampi compared to the MCI and AD. We found a similar pattern comparing cognitively frail group to MCI and Alzheimer’s disease group, although cluster extents were smaller (**Figure 2C-D**).

**Table 2.**
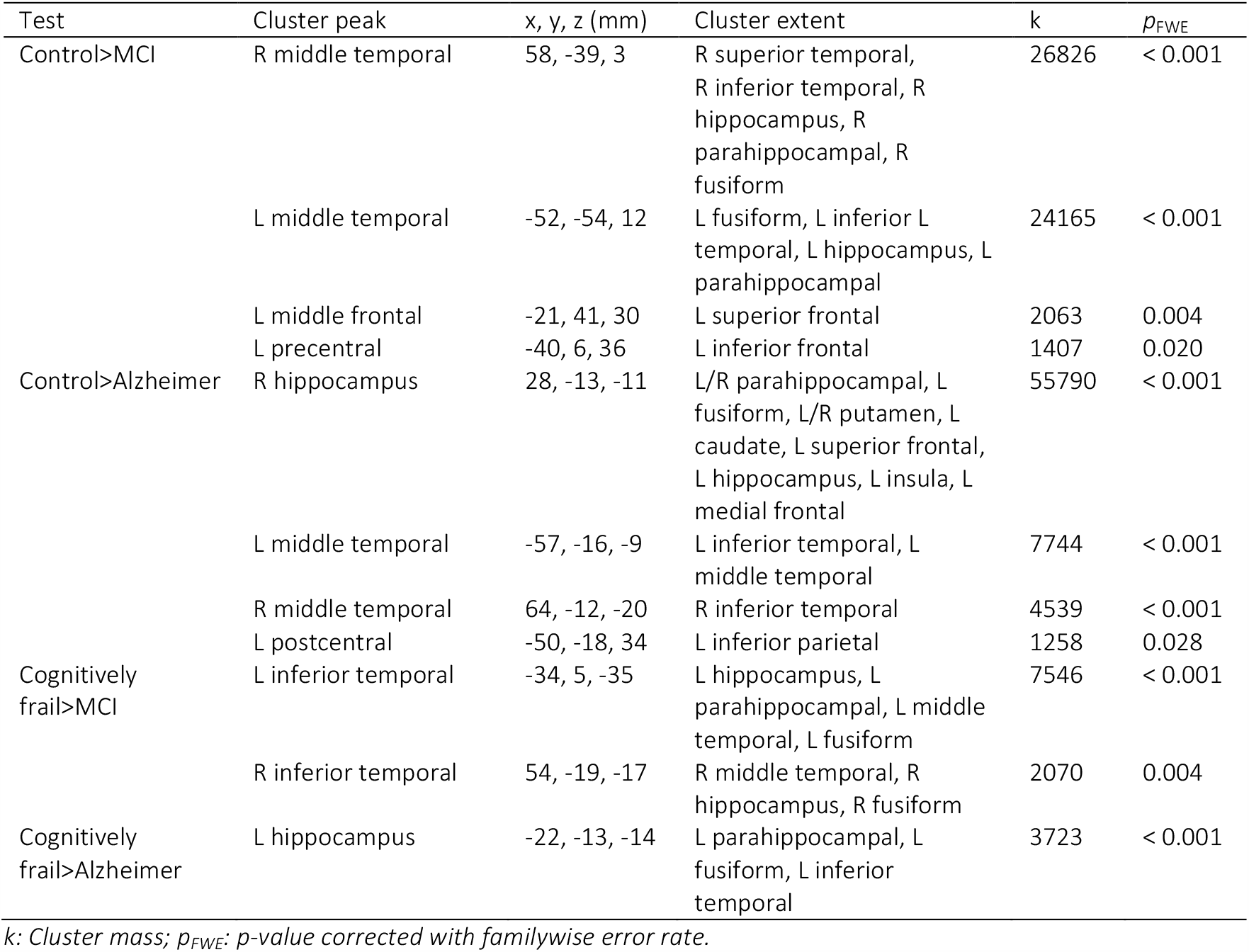
VBM cluster level results. Columns in the table indicate the peak cluster, coordinates of the peak in mm, the extent of the cluster, the cluster mass and corrected p-value for the cluster respectively.

| Test | Cluster peak | x, y, z (mm) | Cluster extent | k | $p_{FWE}$ |
| --- | --- | --- | --- | --- | --- |
| Control>MCI | R middle temporal | 58, -39, 3 | R superior temporal, R inferior temporal, R hippocampus, R parahippocampal, R fusiform | 26826 | < 0.001 |
|  | L middle temporal | -52, -54, 12 | L fusiform, L inferior temporal, L hippocampus, L parahippocampal | 24165 | < 0.001 |
|  | L middle frontal | -21, 41, 30 | L superior frontal | 2063 | 0.004 |
|  | L precentral | -40, 6, 36 | L inferior frontal | 1407 | 0.020 |
| Control>Alzheimer | R hippocampus | 28, -13, -11 | L/R parahippocampal, L fusiform, L/R putamen, L caudate, L superior frontal, L hippocampus, L insula, L medial frontal | 55790 | < 0.001 |
|  | L middle temporal | -57, -16, -9 | L inferior temporal, L middle temporal | 7744 | < 0.001 |
|  | R middle temporal | 64, -12, -20 | R inferior temporal | 4539 | < 0.001 |
|  | L postcentral | -50, -18, 34 | L inferior parietal | 1258 | 0.028 |
| Cognitively frail>MCI | L inferior temporal | -34, 5, -35 | L hippocampus, L parahippocampal, L middle temporal, L fusiform | 7546 | < 0.001 |
|  | R inferior temporal | 54, -19, -17 | R middle temporal, R hippocampus, R fusiform | 2070 | 0.004 |
| Cognitively frail>Alzheimer | L hippocampus | -22, -13, -14 | L parahippocampal, L fusiform, L inferior temporal | 3723 | < 0.001 |

k: Cluster mass; $p_{FWE}$ : p-value corrected with familywise error rate.

### 3.3. Cross-modal mismatch responses

**Figure 3A-D** displays the gradiometer topoplots for each condition in 100 ms time windows across the groups. Following N100, topoplots show a strong burst of bilateral activity in frontal and temporal sensors that is sustained until the end of the epoch. Note that compared to the associative deviant and standard, novelty deviant induced a stronger and more widespread activity across the frontotemporal sensors. The gradiometer topoplots are given here for visualisation only, the topoplots were not tested for task and group effects to avoid double dipping.

**Figure 3.**
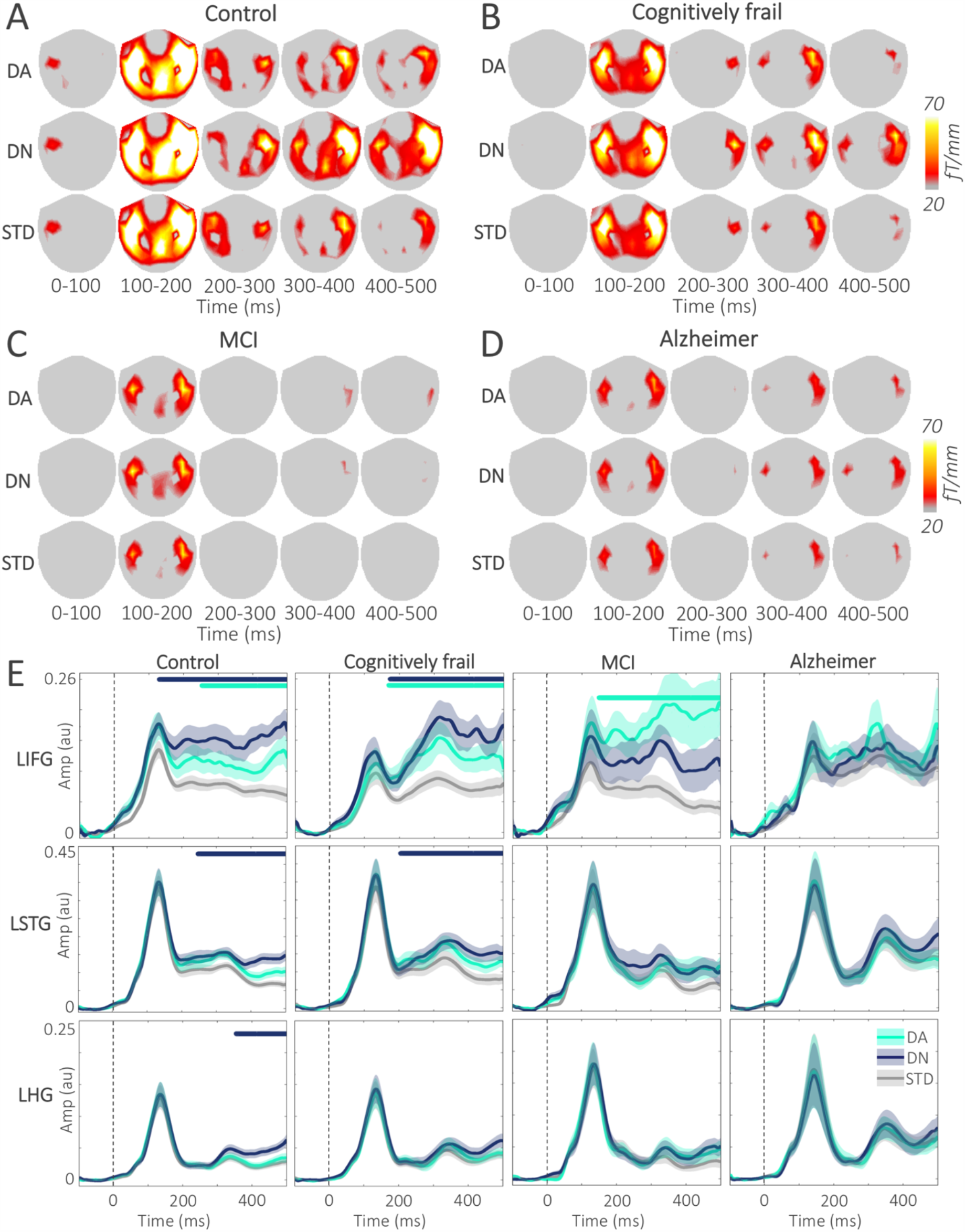
Associative and novelty deviant group means. **A-D**. Topoplots show the mean gradiometer activity across the scalp for the DA, DN and STD conditions in 100 ms time windows in four groups. Note that the DN amplitude is higher after the N100 peak, compared to both DA and STD conditions in control and cognitively frail groups. The gradiometer activity in the MCI and Alzheimer’s disease groups are weaker compared to the control and cognitively frail groups. **E**. Plots display the RMS time series for the left hemisphere ROIs for simplicity. Dashed vertical lines mark the sound onset. Note that the amplitude differences between the deviants and the standard in the frontal regions were larger than the temporal regions, and the deviant effects are stronger in the control and cognitively frail groups; and that there is considerably higher variance in the MCI group. *Amp: Amplitude; AU: Arbitrary units; DA: Associative deviant; DN: Novelty deviant; fT: Femtotesla; LHG: Left Heschl’s gyrus; LIFG: Left inferior frontal gyrus; LSTG: Left superior temporal gyrus; STD: Standard*.

We tested the time series of each deviant with respect to the standard, across 6 ROIs (**Table 3**, **Figure 3E**). We found strongest effects for the novelty deviant in the bilateral IFG early in the epoch, following the onset of the sound. The effects seen in the cognitively frail group mirrored the controls. Further, novelty deviant effects were found across all the ROIs in the control group. MCI and Alzheimer’s disease groups showed no significant novelty effects in the IFG, and weaker clusters limited to STG and HG. Associative deviant effects were found in the IFG across all groups, and in overlapping time windows starting around 200 ms after the sound onset.

**Table 3.** ROI task effects.

| Contrast | ROI | Group | $k$ | $p_{cor}$ | Time (ms) |
| --- | --- | --- | --- | --- | --- |
| STD-DN | LIFG | Control | -1478.03 | < 0.001 | 31-500 |
|  |  | Frail | -938.79 | < 0.001 | 175-500 |
|  | RIFG | Control | -1631.81 | < 0.001 | 1-500 |
|  | LSTG | Control | -715.93 | 0.002 | 255-500 |
|  |  | Frail | -449.99 | 0.021 | 331-500 |
|  | RSTG | Control | -1068.44 | 0.003 | 172-500 |
|  |  | Frail | -870.95 | 0.003 | 197-500 |
|  |  | Alzheimer | -279.53 | 0.041 | 391-500 |
|  | LHG | Controls | -348.23 | 0.034 | 355-500 |
|  | RHG | Controls | -464.34 | 0.021 | 305-500 |
|  |  | Frail | -253.10 | 0.046 | 380-500 |
|  |  | MCI | -261.62 | 0.041 | 350-470 |
| STD-DA | LIFG | Control | -480.41 | 0.023 | 254-500 |
|  |  | Frail | -680.74 | 0.014 | 171-497 |
|  |  | MCI | -974.42 | 0.004 | 150-500 |
|  | RIFG | Control | -944.45 | < 0.001 | 192-500 |
|  |  | Frail | -561.03 | 0.013 | 188-458 |
|  |  | Alzheimer | -929.57 | 0.003 | 162-500 |

DA: Associative deviant; DN: Novelty deviant; $k$ : Cluster mass; LHG: L Heschl's gyrus; LIFG: L inferior frontal gyrus; LSTG: L superior temporal gyrus; $p_{cor}$ : Corrected $p$ -value; RHG: R Heschl's gyrus; RIFG: R inferior frontal gyrus; ROI: Regions of interest; RSTG: R superior temporal gyrus; STD: Standard.

We tested for the interaction effects between the deviant responses (i.e. STD-DA, and STD-DN) and group. There was no significant interaction between the control and cognitively frail group and between MCI and Alzheimer’s disease groups. Whereas, control group showed stronger associative and novelty deviant responses compared to both MCI and Alzheimer’s disease groups (see **Table S3**). Similarly, cognitively frail group showed the same interaction effects against the MCI and Alzheimer’s disease groups.

### 3.4. Clinical and structural correlates of the cross-modal mismatch

To assess how the deviant responses relate to clinical severity, education and medial temporal lobe atrophy, the linear relationships between the E/MEG contrast means at200-500 ms and each predictor variable were tested using general linear models (**Figure 4A**) whilst controlling for differences in age. This revealed strong relationships between the novelty deviant mean in the LHG and RHG with ACER total and ACER-memory subscale scores: the higher the scores on cognitive tests, the more negative (towards normal) the novelty deviant was. A strong negative relationship between the hippocampal and entorhinal volumes and the deviant response was observed for the left hemisphere ROIs, particularly the LHG. This suggests that medial temporal atrophy is associated with a reduced deviant response even though the MMN response arises from extra-hippocampal auditory cortex. This negative relationship was stronger for the novelty deviant compared to the associative deviant. Education showed moderate positive relationships with the associative deviant in the LSTG and RSTG, whereas it showed a negative relationship with the novelty deviant mean in RIFG. Details of the GLM effects are given in **Table S4**.

**Figure 4.**
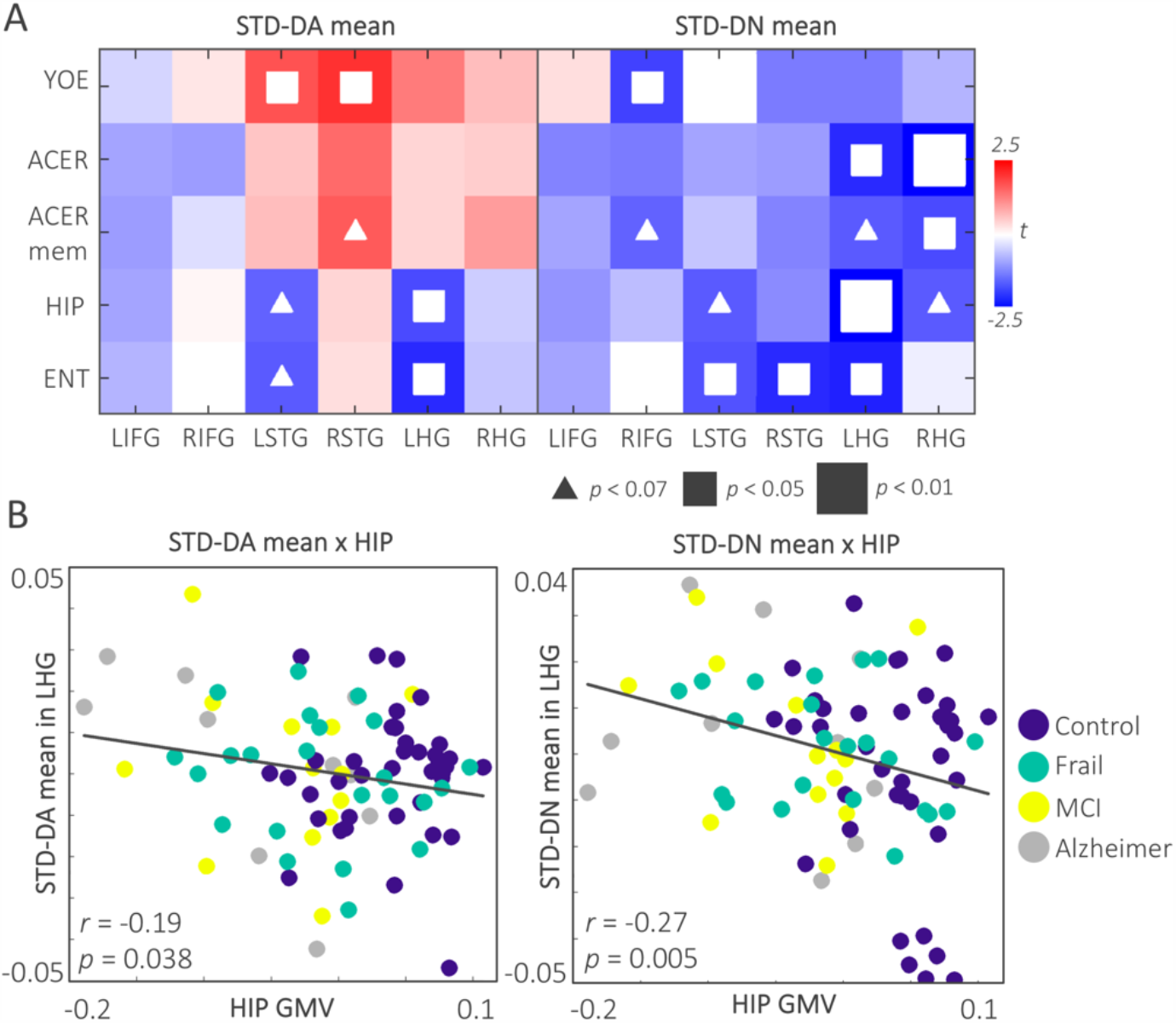
GLM results. **A.** The t-map displays the GLM results across predictors and associative and novelty deviant means for all the ROIs. The white squares indicate significant effects. Note that the effects are stronger for the LHG across the ROIs, and for the novelty deviant compared to the associative deviant. **B.** The scatterplots display the negative relationship between the associative and novelty deviant means in the LHG with the hippocampal GMV across the sample. *ACER: Addenbrooke’s Cognitive Examination Revised; ACER mem: ACER memory subscale; ENT: Entorhinal grey matter volume; GMV: Grey matter volume; HIP: Hippocampal grey matter volume; LHG: L Heschl’s gyrus; LIFG: L inferior frontal gyrus; LSTG: L superior temporal gyrus; RHG: R Heschl’s gyrus; RSTG: R superior temporal gyrus; RIFG: R inferior temporal gyrus; YOE: Years of education*.

We calculated the partial correlations amongst predictor variables correcting for differences in age. Education showed positive correlations with ACER total (*r =* 0.40; *p <* 0.001) and ACER memory subscale scores (*r =* 0.35; *p =* 0.001), but did not correlate with hippocampal and entorhinal volumes. ACER total score correlated with both hippocampal (*r =* 0.53; *p <* 0.001) and entorhinal GMV (*r =* 0.40; *p <* 0.001). Similarly, ACER memory subscale score positively correlated with hippocampal (*r =* 0.55; *p<* 0.001) and entorhinal GMV (*r =* 0.41; *p <* 0.001).

## 4. Discussion

The principal result of this study is that cognitively frail individuals do not resemble people with mild cognitive impairment, in terms of their structural or neurophysiological profile, despite similar levels of underperformance on cognitive tests. The cognitively frail group cannot simply be interpreted as having latent Alzheimer pathology or undiagnosed Alzheimer-related MCI as the cause of their cognitive performance. Population screening using clinical tools (e.g. MMSE, or ACER) is therefore unlikely to selectively identify those with latent Alzheimer’s disease pathology without neuroimaging evidence. There are other associations of cognitive frailty, including lower educational attainment, hearing impairment, and cardiovascular risk factors. Both structural and neurophysiological features of the cognitively frail group were similar to the cognitively more able controls. Structural analyses revealed a higher grey matter volume in the lateral and medial temporal cortices bilaterally in the control and cognitively frail groups compared to mild cognitive impairment and Alzheimer’s disease groups. Like the cognitively healthy controls, the cognitively frail group showed stronger associative and novelty deviant responses compared to MCI or Alzheimer’s disease in relation to hippocampal and entorhinal grey matter volumes.

The cross-modal oddball task was designed to induce deviant responses in the superficial frontotemporal cortex, as neurophysiological markers of hippocampal-dependent associative learning (Chua et al., 2007; Gallo et al., 2004; Giovanello et al., 2004; Konkel et al., 2008; Parra et al., 2009; Sperling, R.A. et al., 2003). We confirmed that the patients with Alzheimer’s disease would show reduced novelty and associative deviant responses. Neurophysiological profiles of the healthy control and cognitively frail groups overlapped, and were significantly stronger compared to the mild cognitive impairment and Alzheimer’s disease groups. The task effects of the novelty deviant responses were observed across all the regions of interest for the control and cognitively frail groups. On the other hand, the group-task interaction effects showing stronger associative and novelty deviant responses for the controls and cognitively frail groups were located in right Heschl’s, bilateral superior temporal and inferior frontal gyri. These results demonstrate that the cognitively frail’s neurophysiological profile is cognitively normal and do not show early neurophysiological signatures of Alzheimer’s disease.

To explore the links between cognitive frailty and Alzheimer’s disease we tested the volumetric differences in the medial temporal lobe, and hippocampal-dependent response to associative learning. Hippocampus is involved in associative memory (Eichenbaum and Lipton, 2008; Giovanello et al., 2004; Jackson and Schacter, 2004; Mayes et al., 2007; Sperling, R. et al., 2003) binding events over time and space (Jackson and Schacter, 2004). Alzheimer’s disease patients show impairments in both sensory and associative memory. They show reduced medial temporal lobe activity to novel scenes parallel to poor performance on explicit memory tests (Düzel et al., 2018; Golby et al., 2005; Rombouts et al., 2000), reduced electrophysiological response to infrequent ‘oddballs’ as measured by the P300 (Daffner et al., 2001; Hedges et al., 2016; Lee et al., 2013) and by mismatch negativity (Engeland et al., 2002; Gaeta et al., 1999; Jiang et al., 2017; Laptinskaya et al., 2018; Mowszowski et al., 2012; Pekkonen et al., 2001; Ruzzoli et al., 2016). Further, they show impairments in multimodal binding, encoding and retrieval of associative memory (Della Sala et al., 2012; Gallo et al., 2004; Parra et al., 2009; Parra et al., 2010; Troyer et al., 2008), reduced hippocampal and entorhinal activity during encoding of novel pairings of stimuli (Dickerson et al., 2005; Sperling, R.A. et al., 2003).

Structurally, early Alzheimer’s disease is characterised by atrophy in the medial temporal lobe as a function of tau burden (Braak et al., 2006; Harper et al., 2017; Hua et al., 2008; Jack et al., 2018; Jak et al., 2007; Mueller et al., 2010; Scheff et al., 2006; Schwarz et al., 2016). Recent studies of cognitive frailty have shown frontotemporal and subcortical atrophy (Del Brutto et al., 2017; Gallucci et al., 2018), white matter hyperintensities (Avila-Funes et al., 2017; Del Brutto et al., 2017; Sugimoto et al., 2019), and decreased white matter microstructure integrity (Avila-Funes et al., 2017). We did not find structural differences between the control and cognitively frail groups in the medial temporal lobe structures, and both groups showed significantly larger hippocampus and entorhinal volumes compared to the MCI and Alzheimer’s disease groups. The cognitively frail individuals do not show early structural signatures of Alzheimer’s disease. The difference between our study and the previous work may lie in the epidemiological approach to baseline recruitment through the population-based Cam-CAN 3000 cohort, rather than clinical referral pathways.

The neuropsychological profile of the cognitively frail resembled mild cognitive impairment group. The cognitively frail scored lower than the healthy controls on every domain of the ACER. Surprisingly, compared to the patients with mild cognitive impairment, they were more impaired on the fluency domain, indicating an executive deficit. Previous studies have suggested that the neuropsychological profile of cognitive frailty differs from mild cognitive impairment in episodic memory with domains of language, visuospatial skills and executive function relatively spared (Collie and Maruff, 2000). The cognitively frailty profile has instead been described in terms of deficits in executive function and attention. The cognitively frail do not use the cues effectively to retrieve stored information (Canevelli et al., 2015; Delrieu et al., 2016); have slower reaction times, make more commission errors on the Sustained Attention to Response Task (O’Halloran et al., 2014; Robertson et al., 2014); and show lower meta-cognitive awareness, error monitoring, disinhibition (Amanzio et al., 2017). The overall cognitive underperformance of the cognitively frail, could be attributed to their lower levels of education and equally to the education bias of the cognitive tests. That is, highly educated individuals perform better on cognitive tests like MMSE and ACER, unless scores are normalised by education (Amaral-Carvalho and Caramelli, 2012; Crane et al., 2006; García-Caballero et al., 2006; Geerlings et al., 1999; Ihle et al., 2018; Jones and Gallo, 2001; Mathuranath et al., 2007; Saliasi et al., 2015; Yassuda et al., 2009).

Our findings support the hypothesis that cognitive frailty represents part of the spectrum of normal neurocognitive function, rather than incipient Alzheimer’s disease. This conclusion calls for a re-evaluation of the prior findings that associate cognitive frailty leads to higher incidence of dementia and cognitive decline (Buchman et al., 2007; Kojima et al., 2016; Rogers et al., 2017; Shimada et al., 2018). These former studies have quantified the dementia incidence including all subtypes of dementia, however this association was highest in non-Alzheimer’s dementias, particularly for vascular dementia (Aguilar-Navarro et al., 2016; Avila-Funes et al., 2012; Gray et al., 2013; Panza et al., 2006; Solfrizzi et al., 2017b). Even though the link between cognitive frailty and Alzheimer’s disease in previous studies was not conclusive, the two clinical entities might share common risk factors such as cardiovascular disease (Frisoli et al., 2015; Fuhrmann et al., 2019; Panza et al., 2006) and hearing impairment (Gates et al., 2002; Panza et al., 2015a; Panza et al., 2015b; Valentijn et al., 2005). Supporting this, the cognitively frail individuals in the Cam-CAN Frail sample showed significantly higher prevalence of hypertension and impaired hearing compared to the controls.

In addition to the cardiovascular risk factors (Fuhrmann et al., 2019; Langlois et al., 2012; Newman et al., 2001; Patrick et al., 2002), the cognitive underperformance of the cognitively frail could be a result of cumulative effects of multiple psychosocial and medical risk factors. Poor nutrition (Chye et al., 2018; Mulero et al., 2011; Rietman et al., 2018), social isolation (Robertson et al., 2013), lower levels of physical exercise (Landi et al., 2010), lack of intellectual cognitive activities (Jung et al., 2010), psychiatric illnesses and long term use of antidepressants (Gray et al., 2015; Paulson and Lichtenberg, 2013), chronic inflammation (Cappola et al., 2003; Solfrizzi et al., 2017a; Walston et al., 2002; Weaver et al., 2002) and lower education levels (Rogers et al., 2017) are known risk factors affecting healthy ageing. In the current study, the cognitively frail group had significantly lower education levels, compared to the healthy controls and MCI. This is a common pattern observed similarly in other frailty studies (Brigola et al., 2019; Margioti et al., 2020). The cognitively frail population have significantly lower occurrence of third-level education (Robertson et al., 2014), and are twice as likely to have no educational qualifications (Rogers et al., 2017). Further, the strong association between levels of education and frailty was linked to mediating socioeconomic, behavioural, and psychosocial factors such as low income, chronic diseases, obesity, depression, unhealthy lifestyle, and chronic stress (Hoogendijk et al., 2014). This is in line with the cognitive reserve hypothesis that an individual’s prior education level and cognitive abilities modify the resilience of their brain structure to disease and injury (Stern, 2002). Longer education in early life and continuing diverse cognitive leisure activities in midlife and old age contribute to an individual’s cognitive reserve, is related to better cognitive functioning in old age (Borgeest et al., 2018; Brigola et al., 2019; Lavrencic et al., 2018; Shafto et al., 2019; Singh-Manoux et al., 2011) and having fewer symptoms of cognitive decline and neuropathology (Chapko et al., 2018; Mortimer et al., 2003; Yoo et al., 2015).

The study has several limitations. Due to the cross-sectional design of the study we are unable to quantify the rates of progression or conversion to dementia from cognitive frailty. Longitudinal cognitive and neuroimaging studies would be able to confirm the rate of conversion to Alzheimer’s disease, and the potential mediators of conversion. Further, this study did not incorporate biomarker testing for Alzheimer’s disease and instead used clinical and neuropsychological criteria to define the groups. The cognitively frail group was defined using an arbitrary threshold on ACER and MMSE test scores. Future studies investigating the link between cognitive frailty and Alzheimer’s disease may test for biomarkers of Alzheimer’s such as tau and amyloid-beta measures acquired from cerebrospinal fluid or positron emission tomography. Future studies can also assess the genetic risk for Alzheimer’s disease using common (e.g. APOE) and less frequent variants associated with the disease, which would help disentangle environmental and psychosocial risk factors from genetic risk factors contributing to the aetiology of cognitive frailty. Further work is needed to clarify the genetic and pathology-based features of cognitive frailty in relation to Alzheimer’s disease and other dementias.

## 5. Conclusions

Our findings provide new evidence that cognitively frail older adults are neurophysiologically and structurally similar to those with successful cognitive ageing, without the hallmarks of mild cognitive impairment despite similarly poor cognitive function. Their underperformance on cognitive tests could be due to lower cognitive reserve and other risk factors across the lifespan.

## Data Availability

Dataset will be made available through the Cam-CAN data portal in the future.

## Abbreviations

ACE-R: Addenbrooke’s Cognitive Examination – Revised
Cam-CAN: Cambridge Centre for Ageing and Neuroscience
DA: Associative deviant
dB: Decibel
DN: Novelty deviant
EEG: Electroencephalogram
ENT: Entorhinal cortex
GMV: Grey matter volume
HIP: Hippocampus
LHG: Left Heschl’s gyrus
LIFG: Left inferior frontal gyrus
LSTG: Left superior temporal gyrus
MCI: Mild cognitive impairment
MEG: Magnetoencephalogram
MMSE: Mini Mental State Examination
MNI: Montreal Neurological Institute
MRI: Magnetic Resonance Imaging
RHG: Right Heschl’s gyrus
RIFG: Right inferior frontal gyrus
RSTG: Right superior temporal gyrus
RMS: Root-mean-square
ROI: Region of interest
STD: Standard
TIV: Total intracranial volume.

## Author’s contributions

JBR, RNH, DN designed the study. LH and RNH designed the paradigm. DN, TE and EK collected the data. RNH preprocessed the structural MRI data. EK pre-processed the MEG data and performed all of the analyses. EK wrote the manuscript, and all authors contributed to the final version.

## Acknowledgements

The Cam-CAN Frail study is funded by the Medical Research Council [SUAG/004 RG91365 & SUAG/046 RG101400]. JBR is supported by the Wellcome Trust [103838] and Medical Research Council [SUAG/004 RG91365]. RNH and TE are supported by the Medical Research Council [SUAG/046 RG101400]. EK is funded by the Dementias Platform UK [MR/L023784/1 & MR/L023784/2], Alzheimer’s Research UK [ARUK-PG2017B-19] and the Holt Fellowship. LH is funded by the Wellcome Trust [103838].

## Supplementary information

**Table S1.** Sample characteristics

|  | Controls | Frail |
| --- | --- | --- |
| Smoking | 58.82% | 52.38% |
| Alcohol use | 43.13% | 19.04% |
| Hypertension | 35.39% | 52.38% |
| Stroke | 0% | 2.38% |
| Heart attack | 1.96% | 4.76% |

**Table S2.** ANCOVA results in cognitive scores

| Test | *F* | *df* | *p* | Control-Frail | Control-MCI | Control – Alzheimer | Frail-MCI | Frail-Alzheimer | MCI-Alzheimer |
| --- | --- | --- | --- | --- | --- | --- | --- | --- | --- |
| *MMSE* | 17.64 | 3,85 | < 0.001 | < 0.001 | 0.010 | < 0.001 | n.s. | 0.006 | 0.002 |
| *ACER* | 55.41 | 3,85 | < 0.001 | < 0.001 | < 0.001 | < 0.001 | n.s. | < 0.001 | < 0.001 |
| *ACER attention* | 9.05 | 3,85 | < 0.001 | 0.0453 | n.s. | < 0.001 | n.s. | 0.027 | 0.028 |
| *ACER memory* | 37.35 | 3,85 | < 0.001 | < 0.001 | < 0.001 | < 0.001 | n.s. | < 0.001 | < 0.001 |
| *ACER fluency* | 13.87 | 3,85 | < 0.001 | < 0.001 | n.s. | < 0.001 | < 0.001 | n.s. | < 0.001 |
| *ACER language* | 8.26 | 3,85 | < 0.001 | 0.008 | 0.042 | < 0.001 | n.s. | n.s. | n.s. |
| *ACER visuospatial* | 11.15 | 3,85 | < 0.001 | 0.001 | n.s. | < 0.001 | n.s. | n.s. | 0.009 |

**Table S3.**
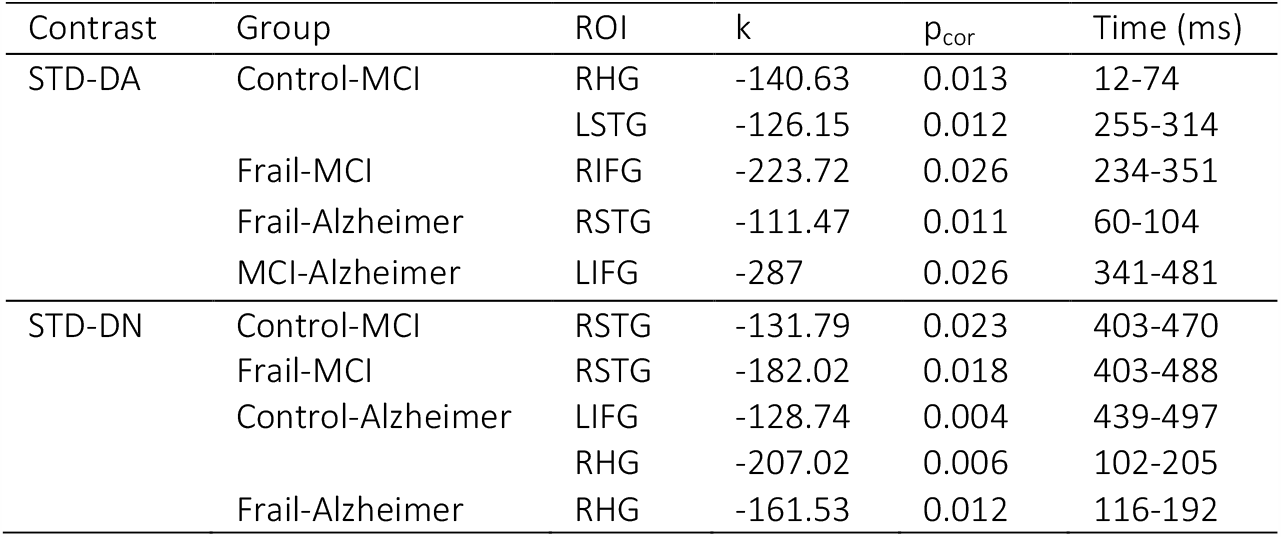
ROI interaction effects

**Table S4.**
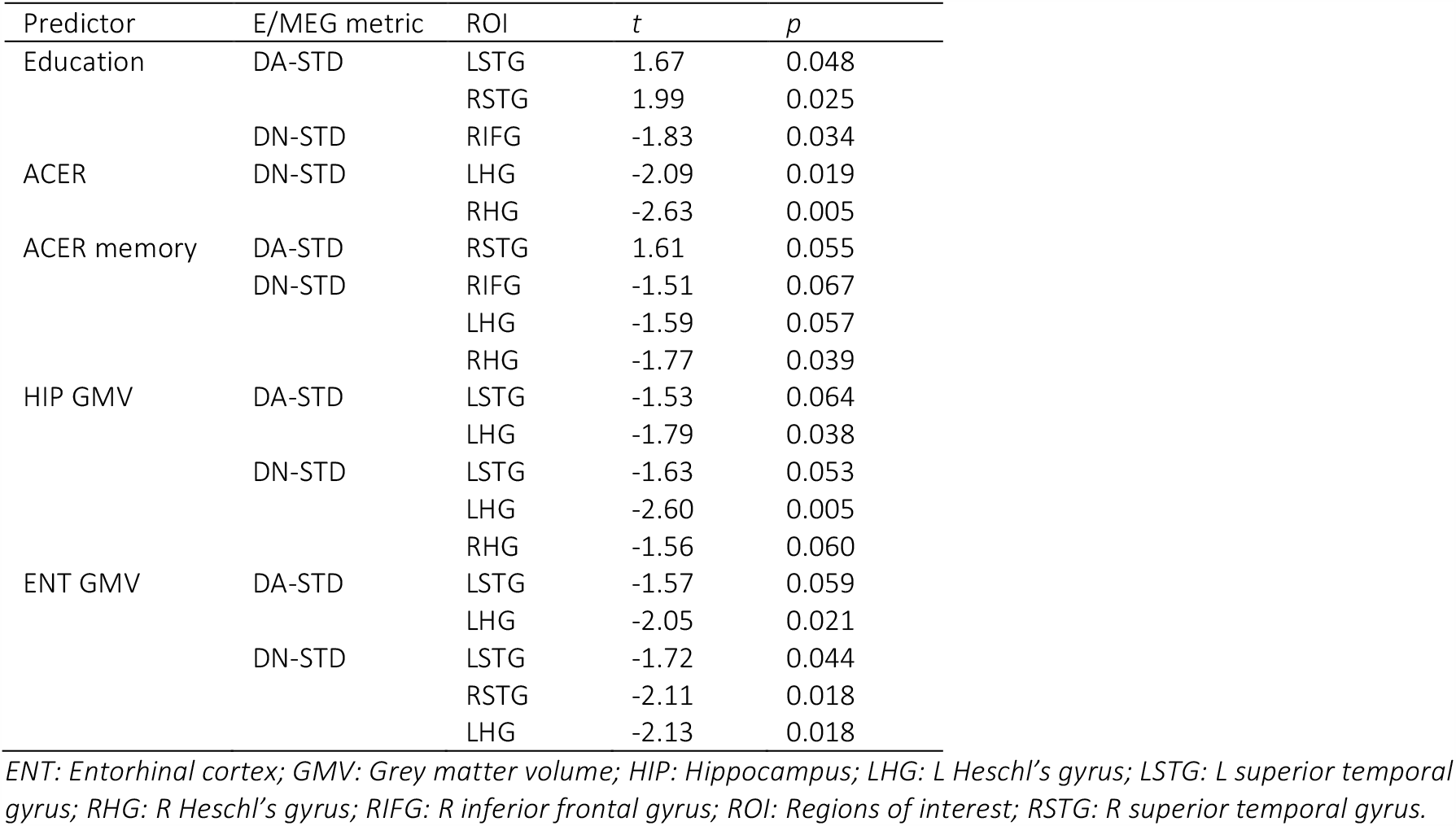
GLM results. Table displays the significant and marginal effects found between the predictors and the DA-STD and DN-STD contrast means in the 200-500 ms time window, after correction for differences in age.

